# AWARENESS AND ACCEPTABILITY OF PRE-MARITAL SCREENING FOR SICKLE CELL AMONG UNIVERSITY STUDENTS IN KENYA

**DOI:** 10.1101/2025.08.21.25334169

**Authors:** Rua sammy, Elijah Ogola, Syokau Ilovi

**Affiliations:** Department of Clinical Medicine and Therapeutics, University of Nairobi

**Keywords:** Premarital Screening, Sickle Cell, Kenya

## Abstract

**Background:** Sickle cell disease is a severe autosomal recessive disorder most prevalent in people of African descent. It is associated with significant morbidity and premature death. Premarital screening for sickle cell is a cost-effective strategy intended to minimize high risk couplings. Premarital screening for sickle cell is yet to be adopted in Kenya.

**Aim:** this study sought to assess awareness and acceptance of premarital screening for sickle cell among a young Kenyan population.

**Materials and Methods:** A descriptive cross-sectional research design utilizing an online, self-administered, questionnaire was conducted among 314 University students selected by multistage sampling technique. Statistical analysis was done using descriptive methods. The following domains were assessed: Students demographics, Knowledge of inheritance patterns, Attitude and perception towards premarital screening for sickle cell and, acceptance of premarital screening. Chi square test was used to determine significance of association between the study variables and the two groups.

**Results:** A total of 314 unmarried students were recruited with 134 (43%) medical students and 180 (57%) non-medical students. Males were 172(55%) and females 142(45%). Majority; 191(61%) were between ages 21∼25. Overall good knowledge was 241(77%). Sickle cell was identified as a heritable disorder by 123(92%) of medical students and 98(54%) non-medical students. Diagnosis of sickle cell via blood test was correctly identified by 121(90%) medical and 92(51%) non-medical students. Premarital screening was identified as beneficial by 124(93%) medical and 136(76%) non-medical students. Majority of non-medical students (54%) had negative attitude towards pre-marital screening compared 10% medical students. Of all respondents, 74(24%) were unwilling to undergo premarital screening for sickle cell. A Total of 44(14%) were unsure whether to undergo screening or not.

**Conclusion:** Sickle cell is the commonest monogenic disorder in Kenya. Kenyan youths demonstrated significant gaps on knowledge on inheritance patterns, attitude and perception towards premarital screening for sickle cell. There was also limited acceptability to undergo individual premarital screening and genetic counselling. Health education on premarital screening and genetic counselling is an important intervention at reducing burden and prevalence of sickle cell disease.

## Introduction

Sickle cell disease (SCD) is the commonest inherited monogenic disorder affecting people of African descent. Symptoms of SCD often present within the first five years of life with frequent admissions throughout their lives. Sickle cell disease is of great public health concern in Africa where outcomes are often poor due to lack of resources, limited access to supportive therapy and absence of gene therapies. (1,2).

Africa accounts for 51% of SCD globally while the rest is distributed across other regions in the world (3). In many African countries, carrier state of sickle cell is 10% to 40% of the population leading to an average of 2% prevalence of SCD amongst the population (4).

It is estimated that over 14,000 thousand children are born with SCD annually in Kenya. This estimates to a carrier frequency of 1 in every 30 Kenyans. Ultimately, the country’s estimated annual economic burden of sickle cell is Kenya shillings 1.6 billion. This translates to 114,000 Kenya shillings per individual per year (5).

Sufficient awareness through improving knowledge, attitude and perception towards SCD in high-burden areas is key to implementing a successful premarital screening program for sickle cell. However, research has shown poor awareness of the pattern of inheritance among many communities (6,7). High incidence of SCD also still persist in regions with readily available and even free of charge government supported screening and counselling programs in the middle east. This has been attributed due to low uptake of premarital screening and genetic counselling services (8). Poor uptake to premarital screening was also demonstrated in regions like Ghana and Nigeria where the populations studied, had a good grasp on knowledge on pattern of inheritance of the disease and, underlying causes of morbidity and mortality (9,10).

Prevention through carrier identification and genetic counselling remains the most realistic approach at reducing the prevalence and impact of sickle cell disease. This would allow better use of limited resources especially in low-income countries where the condition is most prevalent. Assessing awareness and acceptability to pre-marital screening can help identify gaps in knowledge, attitudes and perceptions. This can then inform public health interventions and policies to reduce the prevalence of the disease. By gauging awareness and acceptability, the study can also provide insights into the effectiveness of existing educational campaigns and suggest areas for improvement (11).

### Materials and methods

A descriptive cross-sectional study carried out between 26^th^ June 2024 and 25^th^ July 2024 targeting 314 unmarried students in their last two years of their courses. Participants were randomly selected to fill an online survey which had been adapted from similar study by Omuemu et al in 2013 in Nigeria (11). Study tool was modified by the investigators to suit the Kenyan context.

The medical student’s curriculum had classes on sickle cell disease and genetics testing while, the computer science curriculum did not cover biology and genetics. Therefore, the computer science group was a reflection of the general population.

The rationale for selecting the final two years of study in both groups was because this population group had a higher chance of assuming parental roles either during studying or soon after completing their education.

Krejcie and Morgan sampling formula which was used to derive sample size of 314 from the student population of 1460. The study used a simple random sampling method to select students who were contacted via email that was obtained from the deans’ offices. An email with the questionnaire was sent to the selected 314 students.

In the questionnaire, before the participant was asked to give or deny consent, the researcher clearly explained the purpose of the study, any benefits associated with the study, the voluntary nature and anonymity of participation to the students. The participants were also informed of any risks or discomforts associated with taking part in the study and their choices in terms of freedom to decline participation in the study and, can withdraw at any time without giving reason and without injustice or loss of any benefits.

The researcher also provided contact information of the study staff in case of further clarification or future questions. The study also did not record the student email and therefore the researcher was blinded to which students participated or declined to participate. Decline of consent was identified by the data collecting application tool (goggle form) and recorded.

For the non-responders, more participants were reselected from the remaining total study population and emails sent. This was repeated until the desired minimum sample size was reached. Each selected participant was given a unique study number. Information was sought on social demographic characteristics, knowledge of inheritance pattern, attitude and perception to pre-marital screening. and, acceptance of pre-marital screening. Responses were monitored and recorded in a goggle form. A second mailing was done after a week if there was no response to the first mailing. Non-response was declared at the end of 2 weeks if there was no feedback from the participant.

Statistical analysis was done using descriptive methods. Frequency and percentages were used to calculate respondents with good knowledge. A score of 1 point allocated for each correct choice and zero for wrong choice. Bloom cut off score of >50% = good knowledge. Attitude and perception outcomes were scored on a 5-point Likert scale of strongly agree, agree, undecided, disagree, strongly disagree. A mean score > 30 out of 55 was considered positive attitude. Mean of >35 out of 65 was considered positive perception.

Chi square test (x2) was used to determine significance of association between the study variables and the two groups. P value less than 0.05 was considered statistically significant.

SPSS version 25 was used to conduct all data analysis.

The study preapproval was obtained from University of Nairobi/Kenyatta National Hospital Ethics and Review Committee (protocol number P125/02/2024) and the National Commission for Science Technology and Innovation (approval number 761733)

**Figure 1:**
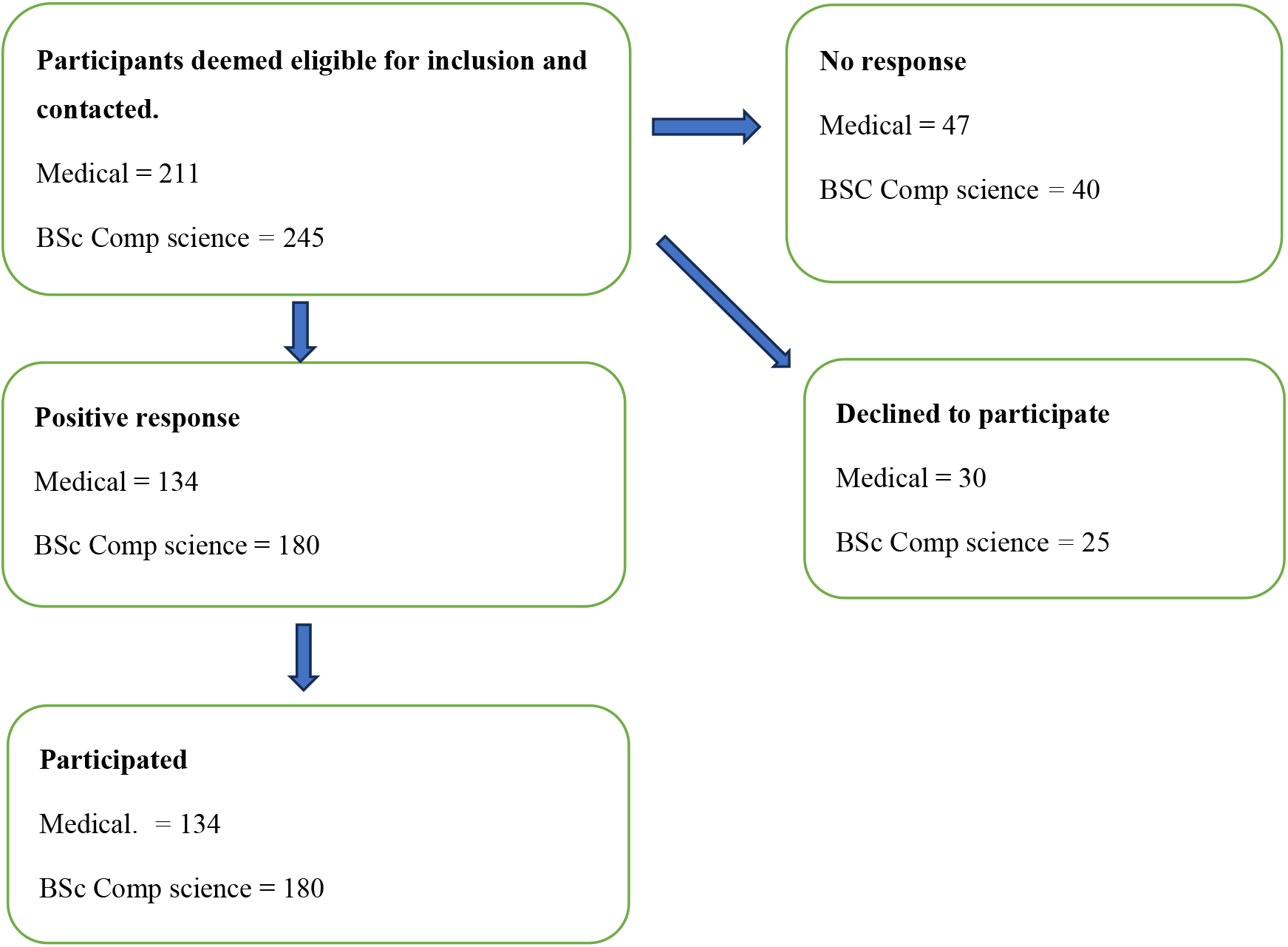
Consort diagram of recruitment process of participants.

## Results

### Social Demographic Data

Age of participants ranged from 20 to 35 years. Majority of students in both groups had no reported relationship with someone who has SCD. Of those who had a positive family history of SCD (20.7%), all respondents indicated they were of distant relationship (non-nuclear or extended family).

**Table 1:**
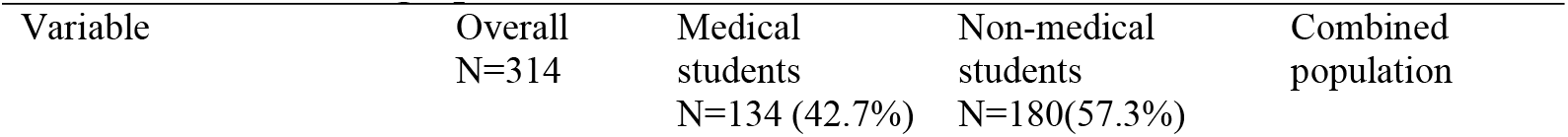
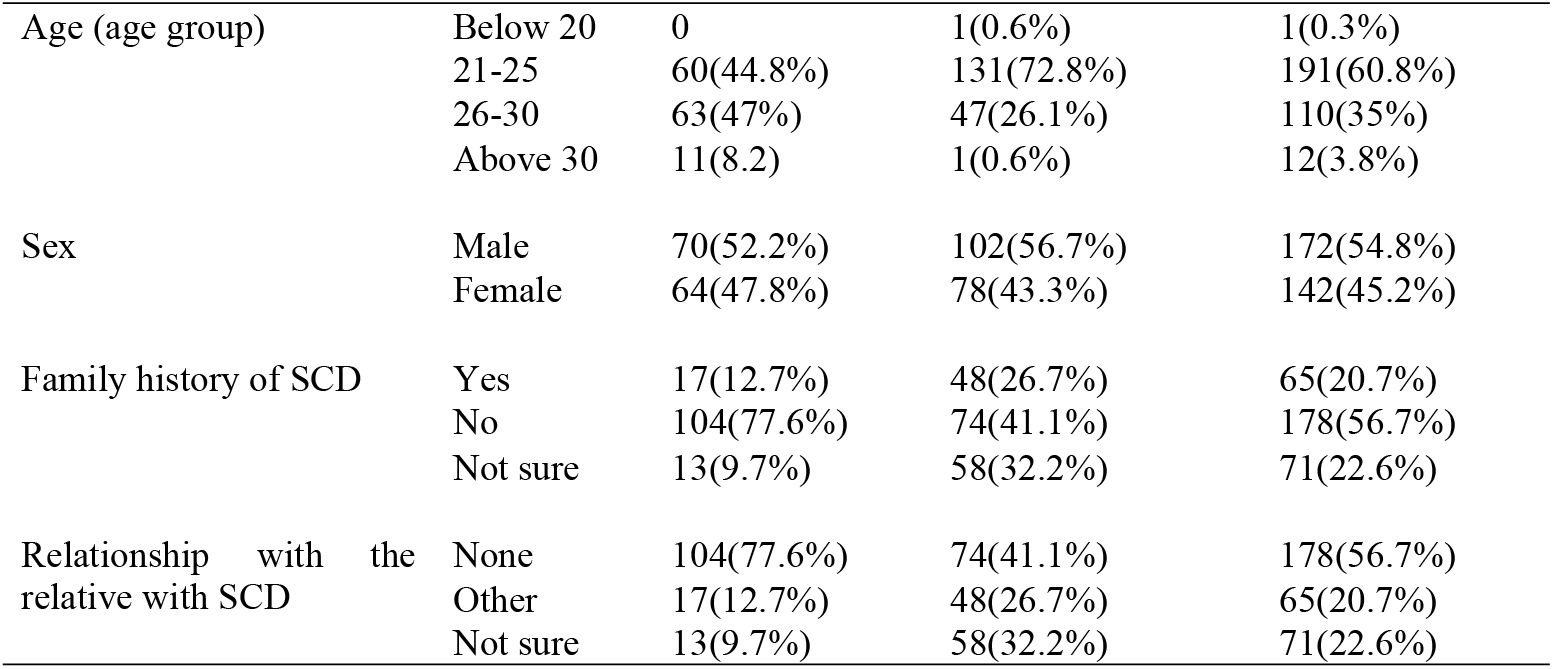
Social Demographic Data.

### Knowledge on Pattern of Inheritance for Sickle Cell Disease among the students

The vast majority of the students (70%) correctly identified that SCD is inherited from parents. The vast majority of medical students (68%) correctly identified a blood test as the primary method for diagnosing SCD. Overall, 78% of students had good knowledge about the best time for a couple to be tested for sickle cell. As shown in the Table, 260 (83%) of the students believed that pre-marital sickle cell screening has benefits.

**Table 2:** Knowledge on Pattern of Inheritance for Sickle Cell Disease.

|  | Course | Good knowledge | Poor Knowledge | P-Value |
| --- | --- | --- | --- | --- |
| Knowledge on SCD definition | Medical | 119(88.8%) | 15(11.2%) | 0.000 |
|  | BSc comp science | 101(56.1%) | 79(43.9%) |  |
|  | Total score | 220(70.1%) | 94(29.9%) |  |
| Knowledge on Probability of having a child with SCD when both parents have the sickle cell trait | Medical | 96(71.6%) | 38(28.4%) | 0.000 |
|  | BSc comp science | 54(30%) | 126(70%) |  |
|  | Total score | 150(47.8%) | 164(52.2%) |  |
| Knowledge on Factors Affecting the Likelihood of a Baby Having Sickle Cell Disease; One parent has the sickle cell | Medical | 94(70.1%) | 40(29.9%) | 0.000 |
|  | BSc comp science | 33(18.3%) | 147(81.7%) |  |
|  | Total score | 127(40.4%) | 187(59.6%) |  |
| Neither parent is carrier or having sickle cell disease | Medical | 119(88.8%) | 15(11.2%) | 0.02 |
|  | BSc comp science | 142(78.9%) | 38(21.1%) |  |
|  | Total score | 261(83.1%) | 53(16.9%) |  |
| Both parents are carriers or having sickle cell disease | Medical | 113(84.3%) | 21(15.7%) | 0.002 |
|  | BSc comp science | 124(68.9%) | 56(31.1%) |  |
|  | Total score | 237(75.5%) | 77(24.5%) |  |
| Knowledge on gender more likely to get sickle cell disease | MBChB | 100(74.6%) | 34(25.4%) | 0.713 |
|  | BSc comp science | 131(72.8) | 49(27.2%) |  |
|  | Total score | 231(73.6%) | 83(26.4%) |  |
| Knowledge on the cause of sickle cell disease- Inherited from parents | Medical | 123(91.8%) | 11(8.2%) | 0.000 |
|  | BSc comp science | 98(54.4%) | 82(45.6%) |  |
|  | Total score | 221(70.4%) | 93 (29.6%) |  |
| Knowledge on diagnosis of sickle cell- Blood test | Medical | 121(90.3%) | 13(9.7%) | 0.000 |
|  | BSc comp science | 92(51.1%) | 88(48.9%) |  |
|  | Total score | 213(67.8%) | 101(32.2%) |  |
| Knowledge on best time for a couple to be tested for sickle cell | Medical | 124(92.5%) | 10(7.5%) | 0.000 |
|  | BSc comp science | 121(67.2%) | 59(32.8%) |  |
|  | Total score | 245(78%) | 69(22%) |  |
| Knowledge on pre-marital sickle cell screening benefits | Medical | 124(92.5%) | 10(7.5%) | 0.000 |
|  | BSc comp science | 136(75.6%) | 44(24.4%) |  |
|  | Total score | 260(82.8%) | 54(17.2%) |  |
|  | TOTAL KNOWLEDGE SCORE | 241(76.6%) | 73(23.4%) |  |

**Figure 2:**
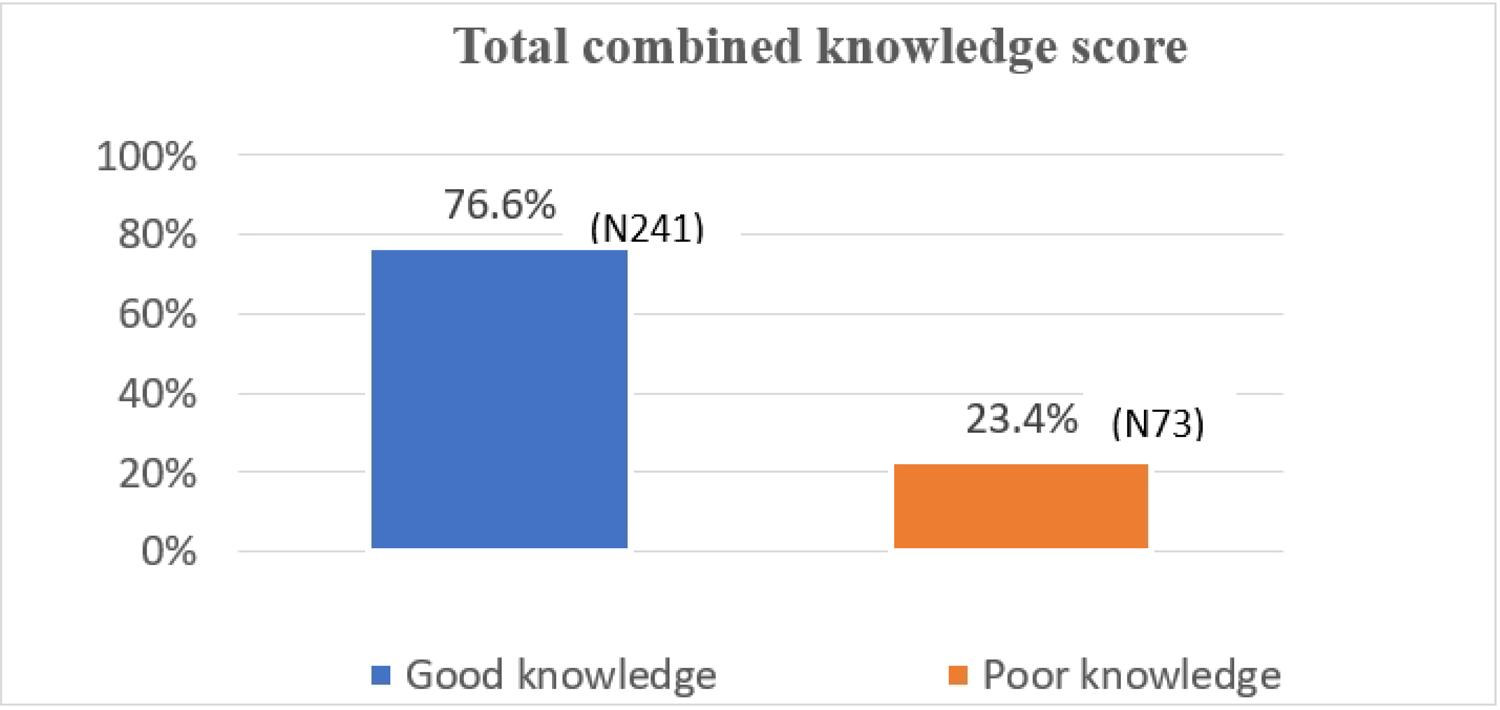
Knowledge score on Pattern of Inheritance for Sickle Cell Disease.

### Attitude towards Pre-Marital Sickle Cell Genotype Screening

Majority of the students 203 (65%) had a positive attitude towards premarital. Most of the medical students (90%) had a positive attitude while majority of computer science students (54%) had negative attitude. The statistically significant result (P-Value 0.000) confirms a clear difference between medical and non-medical students regarding their attitudes towards premarital genotype screening for SCD.

**Table 3:** Attitude score of University of Nairobi students toward sickle cell and premarital screening.

|  |  | **Negative** | **Positive** | **P-Value** |
| --- | --- | --- | --- | --- |
| Course | Medical | 13(9.7%) | 121(90.3%) | 0.000 |
|  | BSc computer science | 98(54.4%) | 82(45.6%) |  |
|  | Total | 111(35.4%) | 203(64.6%) |  |

### Individual Perception on Premarital Sickle Cell Genotype Screening

The majority of the students 227 (72%) had a positive belief or perception about Sickle Cell and Pre-Marital Sickle Cell Genotype Screening. Medical students demonstrated a more positive perception (82%) compared to non-medical 65% (p = 0.001)

**Table 4:** Individual Perception Score of University of Nairobi students on premarital screening.

|  |  | **Positive** | **Negative** | **P-Value** |
| --- | --- | --- | --- | --- |
| Course | Medical | 110(82.1%) | 24(17.9%) | 0.001 |
|  | BSc computer science | 117(65%) | 63(35%) |  |
|  | Total | 227(72.3%) | 87(27.7%) |  |

### Acceptability of Premarital Screening for Sickle Cell

A significant proportion of students was unwilling to be screened with medical at 25 % and, BSc computer science at 23%. Overall acceptance of screening was at 62%.

**Table 5:**
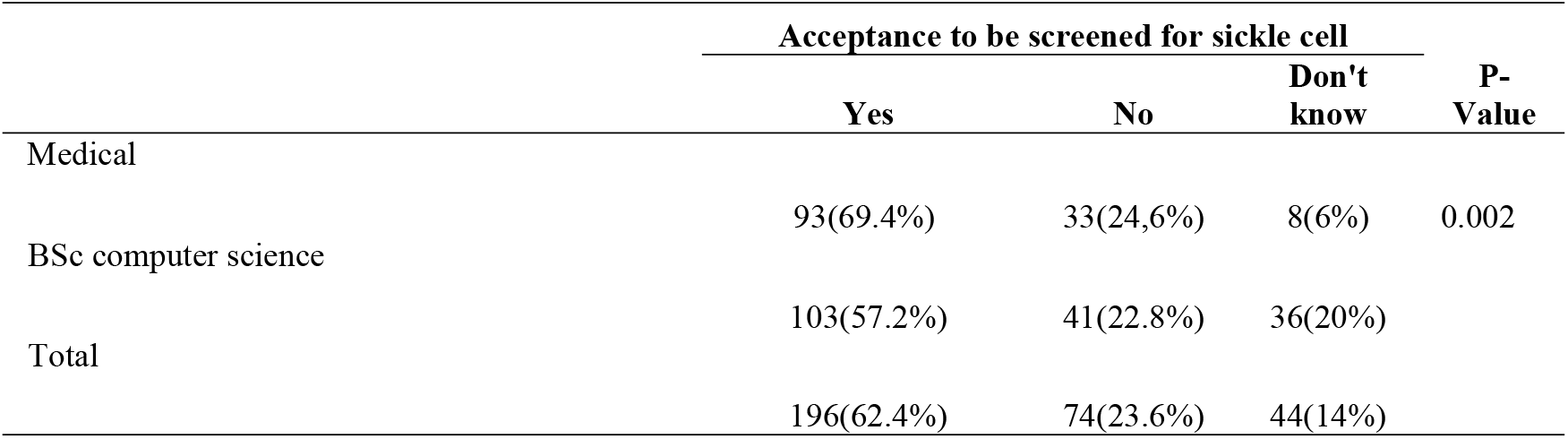
Acceptability of premarital screening among University of Nairobi undergraduate students.

## Discussion

This study sought to determine the level of awareness of inheritance pattern and acceptability of premarital screening for Sickle cell among a young unmarried Kenyan population. We specifically assessed the knowledge, attitudes, perceptions of Sickle cell and, the acceptability of premarital testing among medical and non-medical students. Inclusion of non-medical students provided baseline data on level of knowledge among young lay educated Kenyan youth and inform on appropriate sensitization strategies. Medical students, exposed to the spectrum of sickle disease during their training would provide a useful analysis on whether high level of awareness on sickle cell impact individual uptake of premarital screening.

Majority were 21-25 years age group, comprising over half of the population. A significant portion of the students (57%) were non-medical students compared to the 43% medical students. A majority of the students in both groups did not have a family history of SCD.

Medical students demonstrated superior knowledge across various facets, including the definition of SCD and the likelihood of inheriting the disease from carrier parents. This was so as their curriculum covers SCD, from clinical presentation, testing and inheritance patterns. However, significant number of non-medical students representing the general population in the study were averagely knowledgeable. Many were unaware of the inheritance pattern, diagnosis of SCD, best time for couple to be tested and, on benefits of pre-marital for sickle cell. The non-medical students are a representative sample of real-world awareness of sickle cell prevention strategies and would inform appropriate health intervention strategies.

A similar study by Omuemu et al in 2013, observed high awareness of SCD inheritance pattern and good awareness to premarital screening at 95% at a tertiary Centre in Benin city Nigeria. The majority of respondents also showed good attitude towards premarital screening for sickle cell at an average of 94% across the various social demographic variables. Willingness to undergo screening for sickle cell was at 95%. The high level of awareness and acceptance to premarital screening for sickle cell in that population was attributed to several factors like, mandatory genotype screening at university entry, enlightened population with access to print and electronic media, workshops and lectures and, presence of non-governmental organizations and various interventional measures within the larger community around the institution (12).

Majority of non-medical students had negative attitude towards pre-marital screening at 54%. Individuals with negative attitudes may not see the value and may be less inclined to undergo screening and counseling before marriage. Majority of medical students (90%), demonstrated good attitude towards pre-marital screening. This, aligns with findings from Faremi et al 2018 that, good knowledge on SCD, translates to good attitude towards pre-marital screening and genetic counselling (13).

On perception, we assessed perceived individual susceptibility, perceived benefits and perceived barriers to premarital screening. Negative perception for nonmedical students was higher at 35% while, medical students generally had good perception as only 18% had negative perception.

We identified a notable unwillingness to undergo premarital screening, with 24% of total participants unwilling to undergo screening while 14% unsure whether to undergo screening or not.

Medical students demonstrated a higher acceptance for premarital screening and counselling (69%) as they may have witnessed first-hand the devastating effects of SCD. However, despite this exposure, medical students’ acceptance for individual premarital screening was sub-optimal. This is a pointer to the discordance between knowledge of disease consequences and individual practice. Only 57% of non-medical students were willing to be tested.

Our study findings and implication illustrate considerable gaps in awareness and acceptance to pre-marital screening among Kenyan youth. Creating awareness through media and other channels of communication should be encouraged so that accurate information can reach the community at large. This approach will focus on the most basic level of prevention (primary prevention). Medical personnel can also influence key decision makers in formulation and implementation of strategies and policies that will encourage prevention of SCD as a priority intervention.

## Conclusion

Sickle cell disease is a public health concern in Kenya. Kenyan youths demonstrated significant gaps on knowledge on inheritance patterns, attitude and perception towards premarital screening for sickle cell. There was also limited acceptability to undergo individual premarital screening and genetic counselling. Health education on premarital screening and genetic counselling is an important intervention at reducing burden and prevalence of sickle cell disease.

## Data Availability

All data produced in the present study are available upon reasonable request to the authors

## Recommendations

Given the demographic composition, awareness programs about Sickle Cell Disease should be tailored to the predominant age groups, particularly those aged 18-26 as the median age of first pregnancy in Kenya is 20 years.

Implement educational campaigns that highlight the benefits of pre-marital sickle cell screening.

## Study Limitations

Self-Reported Data therefore responses might be susceptible to social desirability.

Medical and computer science students are considered of high academic standing. Therefore, results could not be generalized.

## Disclosures

This work was completed by the first author in part fulfillment for the degree of Masters of medicine in internal medicine.

No funding was received for this work.

No artificial intelligence software was used for this work.

## BIBLIOGRAPHY

1. Hiran S. Multiorgan dysfunction syndrome in sickle cell disease. The Journal of the Association of Physicians of India. 2005 Jan 1; 53:19–22.

2. Hsu L, Nnodu OE, Brown BJ, Tluway F, King S, Dogara LG, Patil C, Shevkoplyas SS, Lettre G, Cooper RS, Gordeuk VR. White paper: pathways to progress in newborn screening for sickle cell disease in sub-Saharan Africa. Journal of tropical diseases & public health. 2018; 6(2).

3. Piel FB, Hay SI, Gupta S, Weatherall DJ, Williams TN. Global burden of sickle cell anaemia in children under five, 2010–2050: modelling based on demographics, excess mortality, and interventions. PLoS medicine. 2013 Jul 16;10(7):e1001484.

4. Grosse SD, Odame I, Atrash HK, Amendah DD, Piel FB, Williams TN. Sickle cell disease in Africa: a neglected cause of early childhood mortality. American journal of preventive medicine. 2011 Dec 1; 41(6):S398–405.

5. Ministry of Health, “Policy guidelines on infant screening of Sickle Cell Disease.” Accessed: Dec. 14, 2023. [Online]. Available: https://www.health.go.ke/node/581

6. Serjeant GR, Serjeant BE, Mason KP, Gibson F, Gardner R, Warren L, et al. Voluntary premarital screening to prevent sickle cell disease in Jamaica: does it work? Journal of community genetics. 2017 Apr; 8(2):133–9.

7. Smith M, Brownell G. Knowledge, beliefs, attitudes, and behaviors regarding sickle cell disease: Implications for prevention. Social Work in Public Health. 2018 Jul; 33(299-316): 4.

8. Bindhani BK, Davi NK, Nayak JK. Knowledge, awareness, and attitude of premarital screening with special focus on sickle cell disease: a study from Odisha. Journal of community genetics. 2020 Oct; 445–9.

9. Kisakye E, Gavamukulya Y, Barugahare BJ. Sickle cell trait screening in students in a Ugandan university: a cross-sectional study. Journal of International Medical Research. 2022 Nov; 50(11):03000605221138491.

10. Asare EV, Wilson I, Benneh-Akwasi Kuma AA, Dei-Adomakoh Y, Sey F, Olayemi E. Burden of sickle cell disease in Ghana: The Korle-Bu experience. Advances in hematology. Dec 2, 2018.

11. Ango UM, Abiola AO, Yakubu A, Awosan KJ, Yunusa EU. Effect of Health Education Intervention on Knowledge of Sickle Cell Disease and Practice of Voluntary Genotype Couselling and Testing among Students of a Tertiary Institution in Sokoto State, Nigeria. Annals of International Medical and Dental Research. 2018; (4).

12. Omuemu VO, Obarisiagbon OE, Ogboghodo EO. Awareness and acceptability of premarital screening of sickle cell disease among undergraduate students of the University of Benin, Benin City, Edo State. Journal of Medicine and Biomedical Research. 2013 Aug; 12(1): 91–104.

13. Faremi AF, Olatubi IM, Lawal YR. Knowledge of sickle cell disease and pre-marital genotype screening among students of a tertiary educational institution in South Western Nigeria. International Journal of Caring Sciences. 2018 Nov; 11(1): 285–95

